# Efficacy of Combination Treatment for Cervical Precancer Among Women Living with HIV in South Africa: Secondary Outcomes from the ACT 2 Randomized Controlled Trial

**DOI:** 10.64898/2026.03.19.26348810

**Authors:** Carla J. Chibwesha, Nicholas S. Teodoro, Katie R. Mollan, Jessica R. Keys, Chuning Liu, Masangu Mulongo, Sibusisiwe Gumede, Tafadzwa Pasipamire, Mark Faesen, Lisa Rahangdale

## Abstract

**Objective:** We report secondary histologic and high-risk HPV (hrHPV) outcomes from the Acceptability and Feasibility of Combination Treatment for Cervical Precancer Among South African Women Living with HIV (ACT 2) Trial.

**Methods:** We conducted a double-blind Phase 2b feasibility trial of loop electrosurgical excision procedure (LEEP) combined with adjuvant intravaginal 5-fluorouracil (5FU) cream. Women living with HIV (WLWH) and cervical intraepithelial neoplasia (CIN) 2/3 underwent LEEP and were randomly assigned (1:1) to receive 8 doses of 5FU or placebo cream. Our secondary outcomes were (a) regression of cervical disease and (b) clearance of hrHPV.

**Results:** From March 2023 to January 2025, 180 participants underwent LEEP and were randomized to 5FU or placebo cream. Median age was 41 years (IQR: 35-45), 29% had HPV16, 18% had HPV18/45; 99% of women were virologically suppressed (<200 copies/mL) and median CD4 count was 636 cells/uL (IQR: 376-873). 172 participants (95.6%) completed follow-up. At week 24, 96.3% (78/81) in the 5FU group and 82.0% (73/89) in the placebo group regressed to CIN1 or normal histology (PD 14.3%, CI 5.3%, 23.3%). Among participants with positive LEEP margins at week 0, 88.0% (22/25) in the 5FU versus 61.3% (19/31) in the placebo group regressed to CIN1 or normal (PD 26.7%, CI 5.4%, 48.1%). Genotype-specific hrHPV clearance was similar in both groups (5FU: 58.0%, 40/69; Placebo: 53.8%, 43/80; PD 4.2%, CI −11.7%, 20.2%).

**Conclusion:** Clinical outcomes from our Phase 2b trial demonstrates that intravaginal 5FU post-LEEP may be a beneficial adjuvant treatment for CIN2/3.

**Clinical Trial Registration:** NCT05413811

## Introduction

Cervical cancer elimination efforts center around expanding access to Human Papillomavirus (HPV) vaccination and cervical cancer screening. However, treatment of precancerous cervical intraepithelial neoplasia grade 2/3 (CIN2/3) is also a central component to completing the cervical cancer screening cascade. Women living with HIV (WLWH) are at particularly high risk for cervical cancer. Standard treatments are surgical (excision or ablation) but outcomes for WLWH in sub-Saharan Africa are suboptimal, ranging from 20-55% risk of recurrence compared to 5-10% in women without HIV.^1–9^ The need for novel treatment approaches for CIN2/3 in WLWH is therefore substantial, particularly in low- and middle-income countries (LMICs).

Our study was conducted in South Africa, where more than 4 million women are living with HIV and women’s lifetime risk of cervical cancer is among the highest in the world. We previously described ACT 2 (Acceptability and Feasibility of Combination Treatment for Cervical Precancer among South African Women Living with HIV), a randomized placebo-controlled feasibility trial of Loop Electrosurgical Excision Procedure (LEEP) combined with adjuvant self-applied intravaginal 5-flurouracil (5FU).^10^ The trial demonstrated that the combination of LEEP plus adjuvant topical 5-fluorouracil (5FU) is acceptable, safe, and well tolerated. Participants were also highly adherent to the 8-dose 5FU regimen. Here we present clinical outcomes of histologic regression and HPV detection.

## Materials and Methods

Ethical approval was obtained through the University of Witwatersrand Human Research Ethics Committee (201122) and the University of North Carolina Institutional Review Board (20-3565). The trial was registered with the U.S. National Library of Medicine at ClinicalTrials.gov (NCT05413811) and on the South African National Clinical Trials Register. Informed consent was obtained from all participants and instructions were given on use of the study cream. CONSORT guidelines were adhered to during the conduct of this protocol.

The protocol for this trial is previously published.^10^ Briefly, 180 WLWH with CIN 2/3 on LEEP histology were randomized to biweekly (once every two weeks) self-applied 5% intravaginal 5FU cream versus placebo for 8 doses. At 24 weeks post-LEEP, participants underwent colposcopically-directed biopsies and high-risk HPV (hrHPV) testing. The 24-week clinical results (histology and hrHPV) are described herein.

Histology was reviewed at the Bioanalytical Research Corporation (BARC), a South African National Accreditation System (SANAS)-approved clinical trials laboratory in Johannesburg, South Africa. Staining for p16 was used to distinguish between low-grade and high-grade histologic findings as needed by the reviewing pathologist. If multiple biopsies or endocervical curettage was completed, most severe level of histology reported as the final outcome. Discrepancies in findings were resolved by consensus review. All pathologists were blinded to participants’ treatment allocation.

Provider-collected cervical samples (ThinPrep [Hologic, Inc., Marlborough, Massachusetts, USA]) at baseline (week 0), week 4, and week 24 were tested for hrHPV using the GeneXpert system (Cepheid, Sunnyvale, California, USA), which employs a cartridge-based real-time PCR system that reports five separate hrHPV results: (a) HPV16, (b) HPV18/45, (c) HPV 31/33/35/52/58, (d) HPV51/59, (e) HPV39/68/56/66.

### Statistical Analysis

We conducted analyses using the intention-to-treat (ITT) approach, which included all randomized participants. Per-protocol analyses were also performed (not shown). Analyses were conducted in R version 4.4.2 and SAS Enterprise Guide version 8.6.

#### Regression of cervical disease

All eligible participants had CIN2/3 confirmed by LEEP histology measured at study week 0 (baseline for this analysis). The number and percentage of women in each randomization arm with histologic regression to CIN1 or normal at study week 24 were reported with a corresponding 95% confidence interval (CI). A Wilson Score 95% CI was applied for binomial proportions when both the numbers of WLWH with and without histologic regression were ≥3; otherwise, an exact Clopper-Pearson 95% CI was used. Regression of cervical disease was compared between the randomization arms using an estimated difference in proportions and corresponding 95% CI. A Wald 95% CI was used when, in each randomization arm, both the numbers of WLWH with and without regression were ≥3; otherwise, an exact 95% CI was constructed.^11^ The criterion for using the exact method was specified prior to the final analysis. Sample size calculations were based on safety and feasibility primary outcomes described elsewhere.^12^

#### Clearance of hrHPV

The number and percentage of women in each randomization arm with GeneXpert-based detection of genotype-specific hrHPV are reported at weeks 0, 4, and 24. Clearance of HPV was defined as detecting none of the hrHPV types at week 24 that were originally present at week 0. The proportion achieving genotype-specific hrHPV clearance was compared using methods described above for regression of disease.

In an effort to understand the hrHPV response overall and based on individual genotype or genotype-grouping, hrHPV results were analyzed in three ways. First, overall positivity was defined as detection of a positive result for any of the hrHPV genotypes tested. Second, type-specific positivity was evaluated independently for each of the five assay-reported results: HPV 16, HPV 18/45, HPV 31/33/35/52/58, HPV 51/59, and HPV 39/56/66/68. In this non-exclusive analysis, samples positive for multiple genotypes were counted in each respective category. Third, to create mutually exclusive groups for analysis, we derived a hierarchical classification variable. Samples were assigned to the first applicable category based on priority ranking of oncogenic risk: (1) HPV 16; (2) HPV 18/45 (if negative for HPV 16); (3) HPV 31/33/35/52/58 (if negative for HPV 16 and 18/45); (4) HPV 51/59 or HPV 39/56/66/68 (if negative for HPV 16, HPV 18/45 and HPV 31/33/35/52/58); and (5) None.

## Results

Between March 2023 to June 2025, of the 221 WLWH recruited, 210 were eligible for screening. (Figure 1). Of those screened, 27 were ineligible due to either CIN 1 only or cancer diagnosis on LEEP pathology. Ultimately, 180 WLWH underwent randomization, with 90 assigned to each arm. In the placebo and 5FU groups, respectively, 89 and 81 participants used ≥ 6 doses of cream and had 24-week clinical results. Data on acceptability, adherence, safety, and tolerability are reported elsewhere.^12^ Participant baseline demographic information is described in Table 1. The median age of participants was 39 (IQR: 34–44) and 42 (36–46) years, in the 5FU and placebo groups, respectively. Median CD4 count was greater than 600 cells/µL and 99% (176/178) of participants had HIV RNA less than 200 copies/mL; 92% of the participants had viral load less than 40. All participants’ LEEP pathology confirmed CIN 2/3 diagnosis, and cervical HPV was detected in 86% (77/90) of the 5FU and 90% (81/90) of the placebo groups. One person in the placebo group reported HPV vaccination.

**Figure 1.**
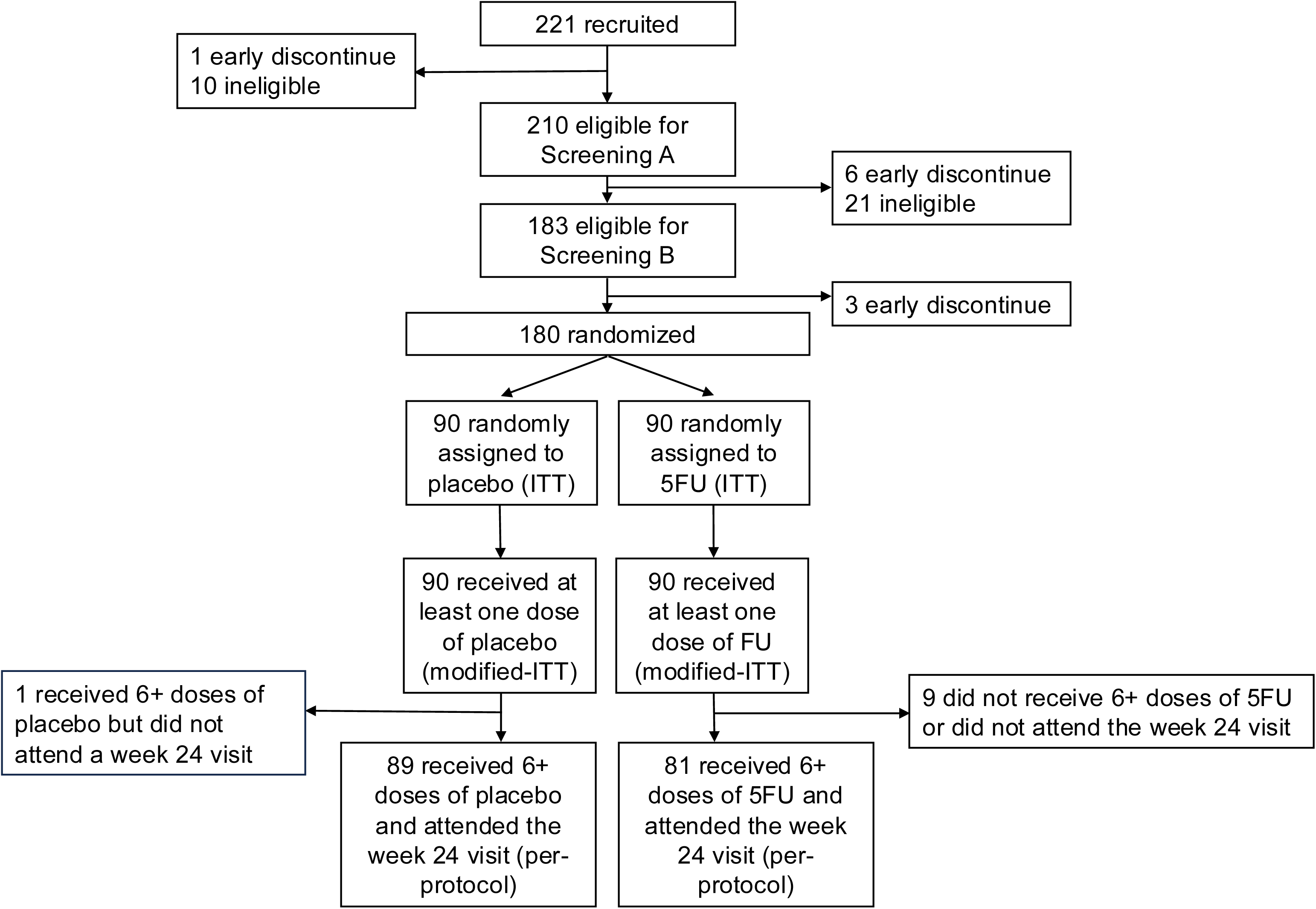
CONSORT Diagram.

**Table 1:** Baseline (Week 0) characteristics of participants.

| **Characteristic**<br>**Median (Q1, Q3) or n (%)** | **Placebo**<br>**n=90** | **5FU**<br>**n=90** | **Overall**<br>**n=180** |
| --- | --- | --- | --- |
| Age at enrollment, years | 42 (36,46) | 39 (34,44) | 41 (35,45) |
| Years since HIV diagnosis | 8 (3,13) | 10 (4,14) | 9 (3,14) |
| CD4+ count, cells/μL | 636 (398, 853) | 632 (342, 898) | 636 (376,873) |
| HIV viral load <200 copies/mL <sup>a</sup> | 88 (99%) | 88 (99%) | 176 (99%) |
| Histology from LEEP |  |  |  |
| CIN 2 | 35 (39%) | 32 (36%) | 67 (37%) |
| CIN 3 | 55 (61%) | 58 (64%) | 113 (63%) |
| HPV (any genotype) | 81 (90%) | 77 (86%) | 158 (88%) |
| HPV Result (hierarchical) |  |  |  |
| HPV 16 | 25 (28%) | 27 (30%) | 52 (29%) |
| HPV 18/45 | 15 (17%) | 17 (19%) | 32 (18%) |
| HPV31/33/35/52/58 | 35 (39%) | 30 (33%) | 65 (36%) |
| HPV 51/59 or 39/68/56/66 | 6 (6.7%) | 3 (3.3%) | 9 (5.0%) |
| None | 9 (10%) | 13 (14%) | 22 (12%) |
| Sexually Transmitted Infections |  |  |  |
| Chlamydia trachomatis | 5 (5.6%) | 7 (7.8%) | 12 (6.7%) |
| Neisseria gonorrhea | 2 (2.2%) | 4 (4.4%) | 6 (3.3%) |
| Treponema pallidum (Syphilis) | 4 (4.4%) | 5 (5.6%) | 9 (5.0%) |
| Trichomonas vaginalis | 10 (11%) | 9 (10%) | 19 (11%) |
| Sexual partners in past year |  |  |  |
| No sexual partners | 18 (20%) | 15 (17%) | 33 (18%) |
| 1 sexual partner | 61 (68%) | 66 (73%) | 127 (71%) |
| 2-5 sexual partners | 10 (11%) | 9 (10%) | 19 (11%) |
| Opted not to answer | 1 (1.1%) | 0 (0%) | 1 (0.6%) |
| Tobacco use, ≥100 times in lifetime | 22 (24%) | 16 (18%) | 38 (21%) |
| Tobacco use in last month | 18 (20%) | 15 (17%) | 33 (18%) |

Cervical Intraepithelial Neoplasia (CIN); Sexually transmitted infections were all treated prior enrollment.<sup>a</sup> Missing data: 2 participants were missing HIV viral load (1 per arm)

At week 24, 96.3% (78/81) in the 5FU group and 82.0% (73/89) in the placebo group regressed to CIN1 or normal histology (per-protocol PD 14.3%, Wald CI 5.3%, 23.3%, sensitivity analysis, exact 95% CI 4.2%, 24.2%). For participants with CIN 2 only diagnosed in the initial LEEP specimen, 96.7 % (29/30) regressed in the 5FU group compared to 85.7% (30/35) in the placebo group. For participants with CIN 3 diagnosed in the LEEP specimen, 96.1% (49/51) regressed in the 5FU group compared to 79.6% (43/54) in the placebo group. No participants progressed to cancer in the 24-week follow up period. In an exploratory analysis of participants with positive LEEP surgical margins, 88.0% (22/25) in the 5FU versus 61.3% (19/31) in the placebo group regressed to CIN1 or normal (PD 26.7%, CI 5.4%, 48.1%).

Overall, at week 0, 87.1% (158/180) participants had at least one hrHPV genotype detected. Post-LEEP, at week 4 and week 24, 47.2% (85/180) and 47.4% (81/171) had hrHPV detected, respectively. HPV detection at week 24 in placebo versus 5FU was similar in both groups (5FU: 58.0%, 40/69; Placebo: 53.8%, 43/80; PD 4.2%, CI −11.7%, 20.2%). See Table 2 and Supplemental Table 1 for detailed categorization of HPV results at week 0, week 4, and week 24.

**Table 2:**
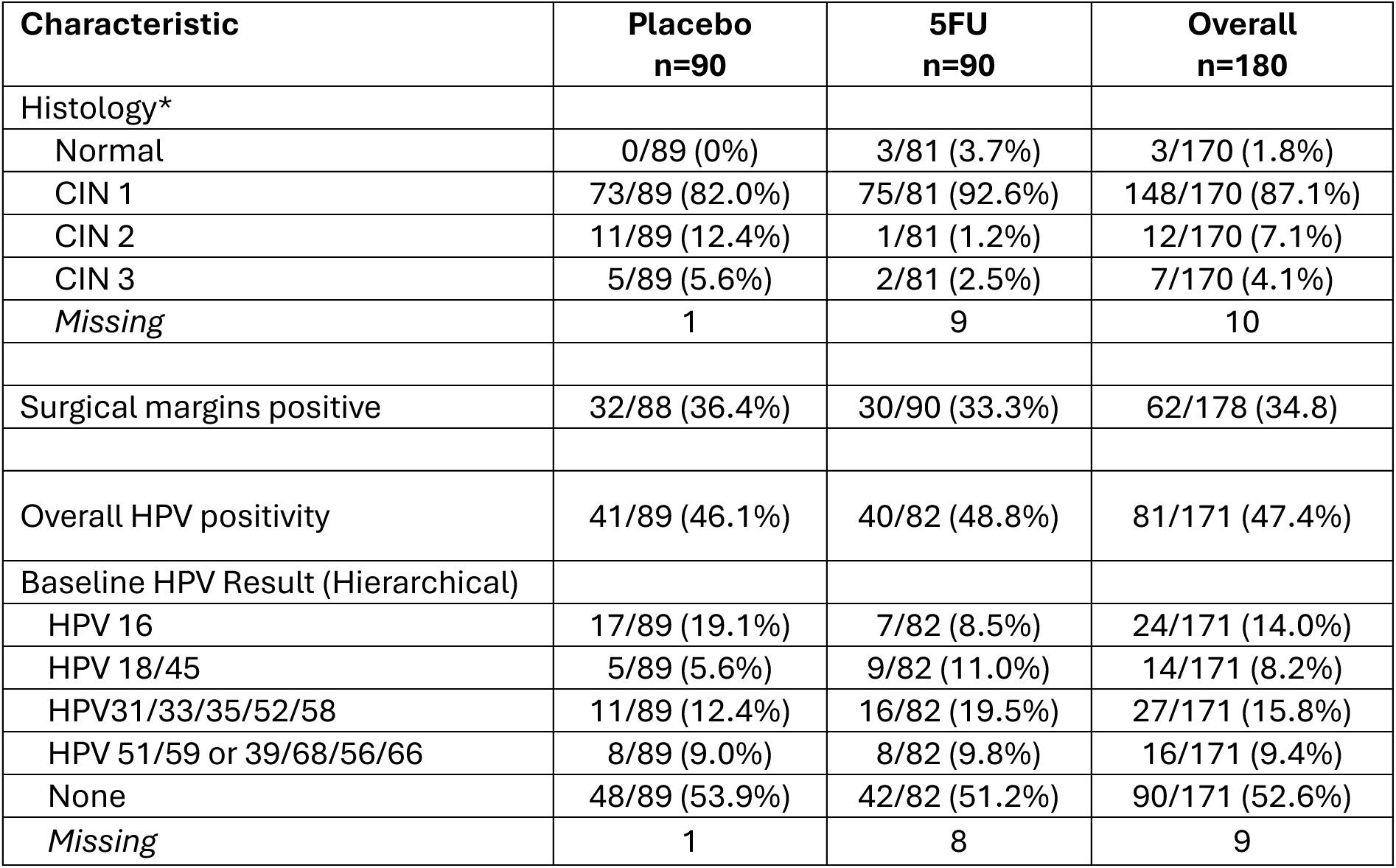
Week 24 post-treatment results.

## Discussion

To our knowledge, this study is the first randomized controlled trial of a topical therapy for treatment of CIN 2/3 in Africa. Our Phase 2b study is also the largest clinical trial of 5FU topical therapy in women, overall, and our primary outcomes indicate acceptability, adherence, safety.^12^ Our secondary clinical outcomes indicate that adjuvant 5FU after LEEP in WLWH may be beneficial to decrease persistent disease at 6 months. Participants in the 5FU group were more likely to have regression of disease to CIN 1 or normal histology overall and were more likely to regress with positive surgical margins. Overall benefit for HPV clearance, was mostly notable at week 4 after LEEP and prior to use of study treatment, indicating that perhaps the benefit of adjuvant 5FU is through clearance of residual disease.

Our planned analysis was based on regression to CIN 1 or normal at 6 months, but if viewed on the basis of risk reduction of persistent CIN 2/3, the probability ratio (PR) is 0.21, demonstrating a 79% relative risk reduction of persistent CIN 2/3 at 6 months post-LEEP. The number needed to treat (NNT) is 7 (95% exact CI 4, 24) indicating that, on average, treating seven women with adjuvant intravaginal 5FU would result in one additional case of regression from CIN2/3 to CIN1/normal histology at week 24.

Prior studies of 5FU have also noted beneficial histologic findings. A U.S. based AIDS Clinical Trial Group study (ACTG 200) conducted before the advent of highly active antiretroviral therapy (HAART) demonstrated that WLWH self-applying intravaginal 5FU every 2 weeks for 6 months as adjuvant therapy to LEEP or cryotherapy were less like to experience CIN2/3 recurrence.^13^ The 101 participants were relatively immunocompromised: 35% had CD4 counts of <200 cells/mm^3^ and half were antiretroviral therapy (ART)-naïve. A more recent study in women without HIV and with CIN 2 demonstrated benefit of primary therapy of using self-applied vaginal 5FU every two weeks for 16 weeks.^14^ Qualitative and pilot studies in Kenya also show safety and acceptability of intravaginal treatment of 5FU after CIN 2/3 diagnosis by both WLWH and male partners^15–18^ A systematic review of commercially available topical therapies determined 5FU to likely be most effective.^19^

Strengths of this study include this data coming from a placebo-blinded randomized controlled trial for a safe, feasible, readily available, self-applied low-cost generic drug.^12^ Data were collected in a systematic manner with high level of participant adherence and retention. WLWH were relatively immunocompetent and taking ART with excellent virologic control. Histology review was blinded and p16 immunohistochemistry was used by pathology, a rare finding in LMIC studies. Lastly, a clinically meaningful result of our study is that adjuvant therapy had benefit in the setting of positive margins which may be a generalizable finding for other countries which perform LEEP for treatment of CIN 2/3.

Limitations of this study include limited sample size due to our Phase 2b feasibility trial not being fully powered for clinical efficacy outcomes. There were more participants in the 5FU group (9) compared to placebo (1) without 24-week histology and HPV results though this study dropout rate was within our anticipated sample size for the primary outcome focused on feasibility. It also is notable that the percentage of trial participants with negative LEEP surgical margins was much higher than in usual clinical care at the study site, which could have impacted the measured benefit of adjuvant therapy. Future Phase 3 studies should include larger sample sizes to definitively answer clinical outcomes.

### Conclusions

Improved treatment options for CIN2/3 are urgently needed for WLWH. Clinical outcomes from our trial demonstrate that intravaginal 5FU post-LEEP has potential as an adjuvant treatment for CIN2/3, particularly in the setting of positive surgical margins. While additional research on clinical outcomes is recommended and a topical treatment which promotes both histologic and virologic clearance would be ideal, findings from our ACT 2 trial demonstrate a strong benefit from 5FU - a treatment with acceptable side effects, low cost, and ready availability across the world – to enhance the cervical cancer prevention toolkit in LMICs where access to vaccination, screening, and surgical treatment is limited.

## Data Availability

Data collection tools and a de-identified dataset are available for research, educational, and non-profit purposes upon request and with appropriate institutional research board and ethics committee approvals.

## Acknowledgements

The authors thank Meghana Mugi, Nhlanhla Kgomari, Tumelo Dlamini, Noluthando Mabandla, Jermina Nkoana, Cassius Dintwe, and Emily Lewis for their invaluable assistance during the trial.

## Author contributions

CJC and LR conceived and designed the project with input from KRM, NST, JRK, MM, and MF. CJC, NST, MM, SL, TP, MM, and MF participated in participant recruitment and data collection. KRM, JRK, and CL planned and conducted the analyses with input from CJC, NST, and LR. CJC and LR drafted the manuscript and coordinated edits. All authors provided critical input during development of the manuscript and approved the final version. Authors are responsible for the correctness of the statements provided in the manuscript.

## Ethics approval

This study was performed in accordance with the principles of the Declaration of Helsinki. We obtained ethical approval from the University of Witwatersrand Human Research Ethics Committee (201122) and the University of North Carolina at Chapel Hill Institutional Review Board (20-3565). Informed consent was obtained from all participants included in the study.

## Conference presentation

The preliminary results of this study were presented at the 37th Annual Conference of the International Papillomavirus Society (IPVC 2025), Bangkok, Thailand, October 2025.

