## Supplemental Table 1 for "Efficacy of Combination Treatment for Cervical Precancer Among Women Living with HIV in South Africa: Secondary Outcomes from the ACT 2 Randomized Controlled Trial"

**Supplemental Table 1: Detection of HPV at Week 0, Week 4, and Week 24 (Intention to treat analysis)**

| | Placebo | | | 5FU | | |
|---|---|---|---|---|---|---|
| | Week 0 (N=90) | Week 4 (N=90) | Week 24 (N=90) | Week 0 (N=90) | Week 4 (N=90) | Week 24 (N=90) |
| **HPV16 Result** | | | | | | |
| Positive | 25 (27.8%) | 17 (18.9%) | 17 (19.1%) | 27 (30.0%) | 11 (12.2%) | 7 (8.5%) |
| Negative | 65 (72.2%) | 73 (81.1%) | 72 (80.9%) | 63 (70.0%) | 79 (87.8%) | 75 (91.5%) |
| Missing | 0 | 0 | 1 | 0 | 0 | 8 |
| **HPV18/45 Result** | | | | | | |
| Positive | 16 (17.8%) | 5 (5.6%) | 8 (9.0%) | 20 (22.2%) | 8 (8.9%) | 9 (11.0%) |
| Negative | 74 (82.2%) | 85 (94.4%) | 81 (91.0%) | 70 (77.8%) | 82 (91.1%) | 73 (89.0%) |
| Missing | 0 | 0 | 1 | 0 | 0 | 8 |
| **HPV31/33/35/52/58 Result** | | | | | | |
| Positive | 52 (57.8%) | 26 (28.9%) | 22 (24.7%) | 51 (56.7%) | 27 (30.0%) | 21 (25.6%) |
| Negative | 38 (42.2%) | 64 (71.1%) | 67 (75.3%) | 39 (43.3%) | 63 (70.0%) | 61 (74.4%) |
| Missing | 0 | 0 | 1 | 0 | 0 | 8 |
| **HPV51/59 Result** | | | | | | |
| Positive | 9 (10.0%) | 6 (6.7%) | 4 (4.5%) | 15 (16.7%) | 11 (12.2%) | 8 (9.8%) |
| Negative | 81 (90.0%) | 84 (93.3%) | 85 (95.5%) | 75 (83.3%) | 79 (87.8%) | 74 (90.2%) |

| | | | | | | |
|---|---|---|---|---|---|---|
| Missing | 0 | 0 | 1 | 0 | 0 | 8 |
| **HPV39/56/66/68 Result** | | | | | | |
| Positive | 20 (22.2%) | 15 (16.7%) | 14 (15.7%) | 16 (17.8%) | 12 (13.3%) | 9 (11.0%) |
| Negative | 70 (77.8%) | 75 (83.3%) | 75 (84.3%) | 74 (82.2%) | 78 (86.7%) | 73 (89.0%) |
| Missing | 0 | 0 | 1 | 0 | 0 | 8 |
| **Overall HPV Result** | | | | | | |
| Positive | 81 (90.0%) | 42 (46.7%) | 41 (46.1%) | 77 (85.6%) | 43 (47.8%) | 40 (48.8%) |
| Negative | 9 (10.0%) | 48 (53.3%) | 48 (53.9%) | 13 (14.4%) | 47 (52.2%) | 42 (51.2%) |
| Missing | 0 | 0 | 1 | 0 | 0 | 8 |
| **Hierarchical hrHPV type** | | | | | | |
| HPV 16 | 25 (27.8%) | 17 (18.9%) | 17 (19.1%) | 27 (30.0%) | 11 (12.2%) | 7 (8.5%) |
| HPV 18/45 (without HPV 16) | 15 (16.7%) | 2 (2.2%) | 5 (5.6%) | 17 (18.9%) | 8 (8.9%) | 9 (11.0%) |
| HPV 31/33/35/52/58 (without HPV 16/18/45) | 35 (38.9%) | 17 (18.9%) | 11 (12.4%) | 30 (33.3%) | 19 (21.1%) | 16 (19.5%) |
| HPV 51/59 or 39/68/56/66 (without HPV16/18/45/31/33/35/52/58) | 6 (6.7%) | 6 (6.7%) | 8 (9.0%) | 3 (3.3%) | 5 (5.6%) | 8 (9.8%) |
| None | 9 (10.0%) | 48 (53.3%) | 48 (53.9%) | 13 (14.4%) | 47 (52.2%) | 42 (51.2%) |
| Missing | 0 | 0 | 1 | 0 | 0 | 8 |
| **2 or more hrHPV types** | | | | | | |

| | | | | | | |
|---|---|---|---|---|---|---|
| Yes | 33 (36.7%) | 17 (18.9%) | 16 (18.0%) | 36 (40.0%) | 17 (18.9%) | 11 (13.4%) |
| No | 57 (63.3%) | 73 (81.1%) | 73 (82.0%) | 54 (60.0%) | 73 (81.1%) | 71 (86.6%) |
| Missing | 0 | 0 | 1 | 0 | 0 | 8 |
| **HPV16 and HPV18/45** | | | | | | |
| Yes | 1 (1.1%) | 3 (3.3%) | 3 (3.4%) | 3 (3.3%) | 0 (0%) | 0 (0%) |
| No | 89 (98.9%) | 87 (96.7%) | 86 (96.6%) | 87 (96.7%) | 90 (100%) | 82 (100%) |
| Missing | 0 | 0 | 1 | 0 | 0 | 8 |
| **2 or more among HPV16, HPV18/45, HPV31/33/35/52/58** | | | | | | |
| Yes | 17 (18.9%) | 10 (11.1%) | 13 (14.6%) | 22 (24.4%) | 8 (8.9%) | 5 (6.1%) |
| No | 73 (81.1%) | 80 (88.9%) | 76 (85.4%) | 68 (75.6%) | 82 (91.1%) | 77 (93.9%) |
| Missing | 0 | 0 | 1 | 0 | 0 | 8 |
| **HPV16 or HPV18/45** | | | | | | |
| Yes | 40 (44.4%) | 19 (21.1%) | 22 (24.7%) | 44 (48.9%) | 19 (21.1%) | 16 (19.5%) |
| No | 50 (55.6%) | 71 (78.9%) | 67 (75.3%) | 46 (51.1%) | 71 (78.9%) | 66 (80.5%) |
| Missing | 0 | 0 | 1 | 0 | 0 | 8 |
